# Reduced Olfactory Bulb Volume Accompanies Olfactory Dysfunction After Mild SARS-CoV-2 Infection

**DOI:** 10.1101/2022.07.24.22277973

**Authors:** Marvin Petersen, Benjamin Becker, Maximilian Schell, Carola Mayer, Felix L. Nägele, Elina Petersen, Raphael Twerenbold, Götz Thomalla, Bastian Cheng, Christian Betz, Anna S. Hoffmann

**Affiliations:** Department of Neurology, University Medical Center Hamburg-Eppendorf, Hamburg, Germany; Department of Otorhinolaryngology and Head and Neck Surgery, University Medical Center Hamburg-Eppendorf, Hamburg, Germany; Population Health Research Department, University Heart and Vascular Center, Hamburg, Germany; Department of Cardiology, University Heart and Vascular Center, Hamburg, Germany; German Center for Cardiovascular Research (DZHK), partner site Hamburg/Kiel/Luebeck, Hamburg, Germany; University Center of Cardiovascular Science, University Heart and Vascular Center, Hamburg, Germany

**Keywords:** COVID-19, MRI, Olfactory bulb, Olfactory dysfunction, SARS-CoV-2

## Abstract

**Background:** Despite its high prevalence, the determinants of smelling impairment in COVID-19 remain not fully understood. In this work, we aimed to examine the association between olfactory bulb volume and the clinical trajectory of COVID-19-related smelling impairment in a large-scale magnetic resonance imaging (MRI) analysis.

**Methodology/Principal:** Data of non-vaccinated COVID-19 convalescents recruited within the framework of the Hamburg City Health Study COVID Program between March and December 2020 were analyzed. On average 8 months after recruitment,, participants underwent MRI and neuropsychological testing as well as a structured questionnaire for olfactory function. Between March and April 2022 olfactory function was assessed at an additional timepoint including quantitative olfactometric testing with Sniffin’ Sticks.

**Results:** This study included 233 individuals recovered from mainly mild to moderate SARS-CoV-2 infections. Longitudinal assessment demonstrated a declining prevalence of olfactory dysfunction from 67.1% at acute infection, 21.0% at baseline examination and 17.5% at follow-up. Participants with post-acute olfactory dysfunction had a significantly lower olfactory bulb volume at scan-time than normally smelling individuals. Olfactory bulb volume predicted olfactometric scores at follow-up. Performance in neuropsychological testing was not significantly associated with the olfactory bulb volume.

**Conclusions:** Our work demonstrates an association of long-term smelling dysfunction and olfactory bulb integrity in a sample of individuals recovered from mainly mild to moderate COVID-19. Collectively, our results highlight olfactory bulb volume as a surrogate marker that may inform diagnosis and guide rehabilitation strategies in COVID-19.

## INTRODUCTION

The coronavirus disease 2019 (COVID-19) pandemic, which is caused by severe acute respiratory syndrome coronavirus 2 (SARS-CoV-2), has affected societies worldwide. Olfactory dysfunction is among the most common symptoms in COVID-19 with a reported prevalence of up to 85%.^(1–8)^ COVID-19 features olfactory dysfunction in varying degrees – e.g., anosmia, hyposmia or parosmia – which occur often before the onset of respiratory symptoms.^(3)^ Compared to other COVID-19-related symptoms like cough, fever or fatigue, olfactory dysfunction proved to be more predictive of SARS-CoV-2 infection.^(9, 10)^ Despite its relevance, the understanding of the pathophysiology of SARS-CoV-2-related olfactory dysfunction is still incomplete.

There is ongoing research in the mechanisms of COVID-19-related olfactory dysfunction. Commonly, anosmia in the absence of rhinorrhea or nasal congestion is described as an early symptom which suggests other causal mechanisms than a common cold with conductive deficits causing olfactory dysfunction.^(3, 4, 11, 12)^ SARS-CoV-2 might affect the olfactory system at different breakpoints of its trajectory ranging from disruption of sustentacular cells and olfactory sensory neurons situated in the olfactory mucosa to functional disarray of the olfactory cortex.^(13, 14)^ Yet, there is only vague understanding of how these aspects relate to clinical outcomes.

Magnetic resonance imaging (MRI) provides a promising avenue to investigate the pathomechanistic substrates of olfactory dysfunction in SARS-CoV-2 infection in vivo. Previous MRI studies put emphasis on the integrity of the olfactory bulb (OB) as a structural correlate of olfactory function in general.^(15–17)^ Volume reduction of the OB accompanies olfactory loss in conditions like acute or chronic rhinosinusitis and head trauma.^(18)^ Abnormalities in psychophysical olfactory testing are demonstrably associated with OB volume alterations in health and disease.^(19–21)^ Furthermore, the duration and degree of olfactory loss is proportional to the OB volume.^(22)^ So far, studies relating OB volumetry and olfactory function in COVID-19 rely on case reports and small sample sizes yielding heterogeneous results.^(14, 23–27)^ Therefore, further investigations are warranted.

Based on previous evidence, we hypothesized that olfactory dysfunction in COVID-19 corresponds with interindividual volumetric differences of the OB. To address this hypothesis, we quantified the OB volume based on structural MRI to see if there is a difference in the OB volume between the smell impaired and the normal individuals and performed a longitudinal assessment of olfactory function in a large sample of individuals recovered from mainly mild to moderate COVID-19. We also addressed if the OB volume is predictive for the long-term olfactory function. As affections of the olfactory system might precede COVID-19-related neuropathology, we additionally probed for a relationship of OB alterations and neuropsychological test score results in an exploratory analysis. With this work we aimed to further the understanding of the effects SARS-CoV-2 exerts on the olfactory system and deepen our insight in the longitudinal trajectory of subjective smelling impairment as well as the pathophysiology underlying the clinical sequelae of COVID-19.

## MATERIALS AND METHODS

### Study population and clinical examination

In this work we investigated data from participants of the Hamburg City Health Study (HCHS) Covid Program with available MRI data. A detailed description of the study design has been published separately.^(28, 29)^ Our reporting complies with the Strengthening the Reporting of Observational Studies in Epidemiology (STROBE) statement guidelines.^(30)^ In brief, citizens of the city of Hamburg, Germany, were considered for enrollment if they met two criteria: (1) a laboratory-confirmed positive polymerase chain reaction (PCR) test for SARS-CoV-2, which was obtained between 1^st^ March and 31^st^ December 2020 but at least 4 months prior to study enrollment; (2) age between 45 and 74 years at the time of inclusion. An invitation was issued upon identification via the clinical information system of the University Medical Center Hamburg-Eppendorf or a response to a public call for participation. Recruited participants underwent an extended study protocol of the Hamburg City Health Study (HCHS): besides the standard HCHS work up including MRI and assessment of cognitive function (Trail Making Test B, Word List Recall, Animal Naming Test, Mini Mental State Exam), depressive symptoms (PHQ-9) and quality of life (EQ-5D), participants were required to retrospectively report on disease severity and SARS-CoV-2-associated symptoms via a structured questionnaire.^(31)^ A mild to moderate COVID-19 severity was defined as a symptomatic disease course not requiring intensive care unit treatment. The presented study was only conducted based on the post-SARS-CoV-2 cohort – i.e., the matched cohort of control subjects as described previously could not be leveraged as the required high-resolution T2-weighted MRI data was not available.^(28, 32)^ To assess the trajectory of olfactory function, participants were reinvited between 15^th^ March and 15^th^ April 2022. For a timeline of this study refer to supplementary figure E1. Follow-up investigations comprised a structured questionnaire regarding olfactory function as well as olfactometric assessment via Sniffin’ Sticks Screening 12 test by two trained otorhinolaryngologists (B.B., A.S.H.).^(33, 34)^ The test score ranges from 0 to 12 (0-6: anosmia, 7-10: hyposmia, 11-12: normosmia) and is based on normative information derived from more than 1200 patients assessed with Sniffin’ Sticks Screening and olfactive evoked potentials. Eventually, information about self-reported olfactory dysfunction (based on a yes/no-question) from structured questionnaires was available for three timepoints: (1) during the acute infection, (2) at the baseline investigation and (3) at follow-up. MRI and neuropsychological testing were only performed during baseline examination.

### Ethics approval

Written informed consent was obtained from all participants. The study was approved by the local ethics committee of the Landesärztekammer Hamburg (State of Hamburg Chamber of Medical Practitioners, PV5131) and conducted complying with the Declaration of Helsinki.^(35)^

### MRI acquisition

High-resolution 3D T2-weighted images were acquired at baseline on a 3T scanner (MAGNETOM™Skyra, Siemens Healthineers, Erlangen, Germany) with the following sequence parameters: TR=3200 ms, TE=407 ms, 256 axial slices, ST=0.94 mm, and IPR=0.9×0.9mm.

### Olfactory bulb segmentation

We performed OB segmentation on high-resolution T2-weighted images leveraging a novel fully-automated deep learning-based pipeline specifically designed for OB volumetry.^(36)^ All resulting segmentations underwent visual quality assessment. Exemplary segmentation results are illustrated in *figure 1* as well as *supplementary materials E2-5*. The summed volume of both OBs was used for further analysis.

**Figure 1:**
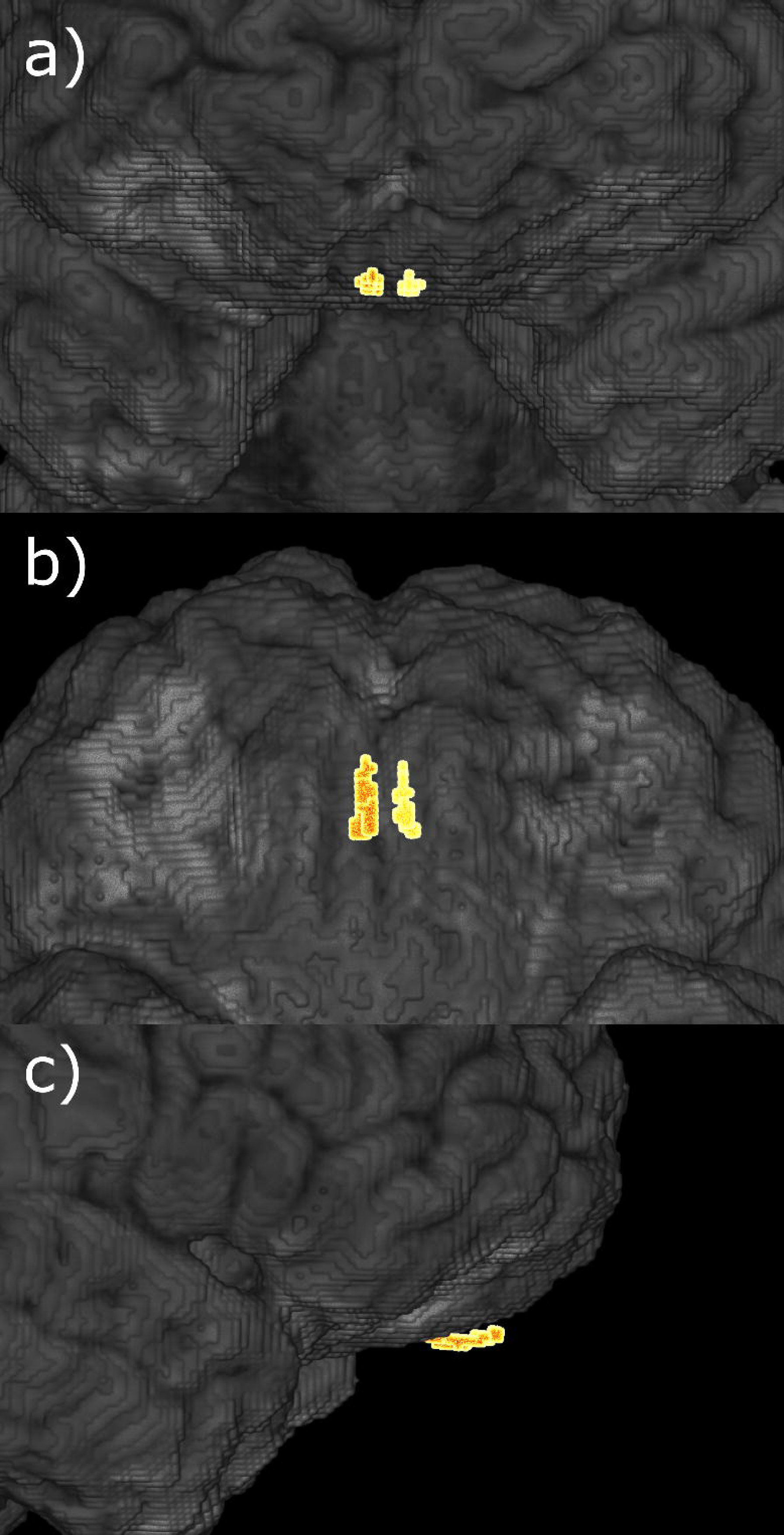
Exemplary 3D visualization of olfactory bulb segmentation results. Volumetric visualization of the left and right olfactory bulb (highlighted) and surrounding brain areas. a) coronal, anterior-posterior; b) axial, inferior-superior; c) sagittal, right-left.

### Statistical analysis

OB volume and olfactometry scores were compared between individuals with and without self-reported olfactory dysfunction (yes/no) at different timepoints employing analyses of covariance (ANCOVA). Multiple linear regression analysis was performed for assessing the linear relationship of olfactometry scores, as well as neuropsychiatric scores with OB volume. A further ANCOVA was performed to test whether the OB volume at baseline differed between individuals with sustained olfactory dysfunction at follow-up and those that recovered until then. The association between the time interval from positive PCR to examination was assessed via Spearman correlation. Age, sex and smoking behavior were included as covariates in ANCOVAs and linear models as they represent potential confounders.^(37)^ Statistical computations and plotting were performed in Python 3.9.7 harnessing matplotlib (v.3.5.1), numpy (v.1.22.3), pandas (v.1.4.2), pingouin (v.0.5.1) and seaborn (v.0.11.2).^(38–42)^

## RESULTS

### Sample characteristics

Data from 233 HCHS Covid Program participants was available for primary analysis. Nine subjects were excluded pre-analysis: 3 since they reported to have had olfactory dysfunction before their SARS-CoV-2 infection, 3 because of a missing OB and 3 because of erroneous segmentations. Thus, data from n=224 participants were available for the final analysis. Sample characteristics are summarized in *table 1*. On average participants were 55.79 [95% confidence interval (CI95); 54.83, 56.73] years old, 44.2% were female and 5.4% were current smokers. 8.4% of participants were hospitalized due to COVID-19.

**Table 1.** Characteristics of Post-SARS-CoV-2 Individuals.

| Demographics |  |
| --- | --- |
| Age in years, mean [CI95] (n) | 55.79 [54.83, 56.73] (224) |
| Female sex at birth, % (n) | 44.2 (224) |
| Education in years, mean [CI95] (n) | 15.83 [15.50, 16.16] (224) |
| Current smokers, % (n) | 5.4 (224) |
| Allergic rhinitis, % | 33.3 (224) |
| Diabetes, % | 4.4 (224) |
| COVID-19 |  |
| Days between first positive SARS-CoV-2 PCR test and baseline (T1), mean [CI95] (n) | 253.15 [242.00, 264.29] (223) |
| Days between first positive SARS-CoV-2 PCR test and follow-up (T2), mean [CI95] (n) | 663.51 [646.04, 680.98] (143) |
| Hospitalization, % | 8.4 (224) |
| Olfaction and olfactory bulb volume |  |
| Subjective olfactory dysfunction at acute infection, % | 67.1 (143) |
| Olfactory dysfunction severity (VAS), acute infection, mean [CI95] (n) | 7.62 [7.06, 8.18] (95) |
| Subjective olfactory dysfunction, T1, % (n) | 21.0 (143) |
| Olfactory dysfunction severity (VAS), T1, mean [CI95] (n) | 3.79 [3.12, 4.47] (29) |
| Subjective olfactory impairment, T2, % (n) | 17.5 (143) |
| Olfactory dysfunction severity (VAS), T2 mean [CI95] (n) | 2.96 [2.07, 3.84] (24) |
| Olfactometry score, T2, mean [CI95] (n) | 10.18 [9.89, 10.48] (143) |
| Olfactory bulb volume in mm<sup>3</sup>, T1, mean [CI95] (n) | 45.53 [43.84, 47.21] (224) |
| Neuropsychological scores |  |
| Trail Making Test B, T1, mean [CI95] (n) | 69.96 [66.55, 73.37] (206) |
| Word List Recall Test, T1, mean [CI95] (n) | 8.51 [8.28, 8.74] (205) |
| Animal Naming Test, T1, mean [CI95] (n) | 28.25 [27.42, 29.08] (206) |
| Mini Mental State Exam, T1, mean [CI95] (n) | 28.42 [28.25, 28.60] (205) |
| Patient Health Questionnaire 9, T1, mean [CI95] (n) | 3.54 [3.05, 4.03] (218) |
| EQ5D, T1, mean [CI95] (n) | 80.74 [78.76, 82.72] (206) |

*Abbreviations:* CI95 = 95% confidence interval of the mean, n = number of data
points, PCR = polymerase chain reaction, post-SARS-CoV-2 = individuals who have
recovered from a severe acute respiratory coronavirus type 2 infection, T1 = at
baseline, T2 = at follow-up.

### Longitudinal trajectory of olfactory function

The baseline examination happened on average 253 days after the positive PCR test prompting recruitment, the follow-up at 663 days. 143 participants were available at follow-up (81 study drop-outs). Of these, 67.1% (n=96) described olfactory dysfunction during the acute phase of infection, 21.0% (n=30) at the baseline examination and 17.5% (n=25) at follow-up (*figure 2a*). Assessment of olfactory dysfunction severity via the visual analogue scale resulted in a coherent trajectory: 7.62 [CI95; 7.06, 8.18] (acute infection), 3.79 [3.12, 4.47] (baseline), 2.96 [2.07, 3.84] (follow-up).

**Figure 2:**
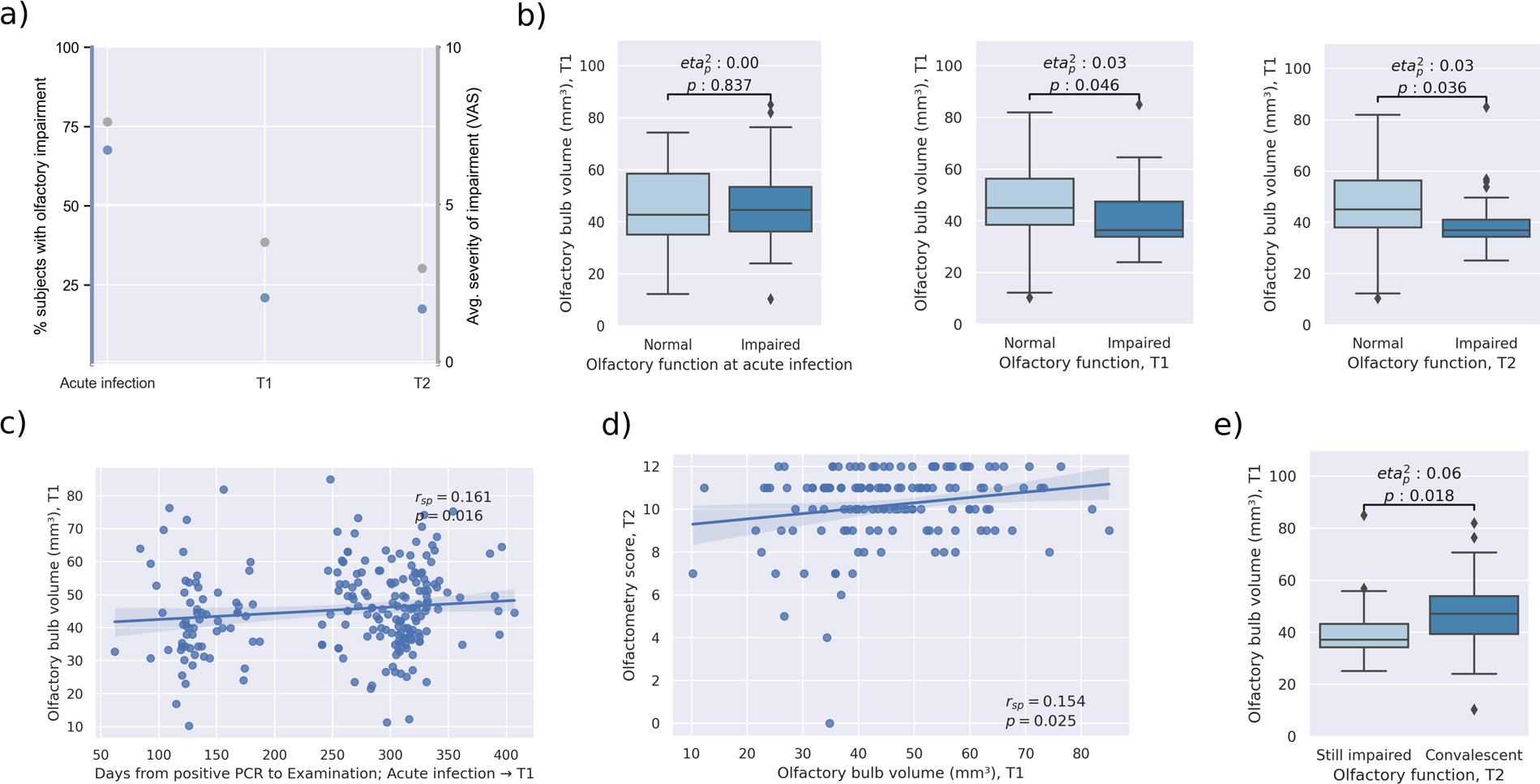
Association of olfactory bulb volume and olfactory function. a) Trajectory of olfactory function along timepoints. Blue dots indicate proportion of individuals with olfactory dysfunction. Gray dots show the average impairment as operationalized by the visual analogue scale. b) Group differences of olfactory bulb volume at baseline between participants with and without olfactory dysfunction with respect to different timepoints. Olfactory bulb volume at baseline was significantly lower in individuals that exhibited olfactory dysfunction during both examination timepoints but not during the acute infection. c) Association of the time interval from positive PCR to examination and olfactory bulb volume. A smaller interval was significantly associated with lower olfactory bulb volume. d) Linear associations between olfactory bulb volume and olfactometry scores. A low olfactory bulb volume at baseline was significantly associated with a lower olfactometry score at follow-up. e) Group differences of olfactory bulb volume between participants with sustained olfactory dysfunction at follow-up and those with recovered olfaction to that point. Olfactory bulb volume was significantly lower in participants with sustained olfactory dysfunction. Abbreviations: 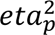 = partial eta squared indicating the effect size as provided by the analysis of covariance, p = p-value, *r*_*sp*_ = spearman correlation coefficient, PCR = polymerase chain reaction, T1 = at baseline, T2 = at follow-up.

### Olfactometry

To assess long term olfactory outcomes, olfactometry with the Sniffin’ Sticks Screening 12 test was performed on participants at follow-up. Olfactometry scores at follow-up were 10.18 [CI95, 9.89, 10.48]. Participants with self-reported olfactory dysfunction at the acute infection did not differ from normally smelling participants regarding olfactometry scores at follow-up (mean [CI95], 10.04 [9.65, 10.44] vs. 10.46 [10.08,10.84], 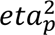=0.01, *p*=0.211; *supplementary materials E6*). Yet, participants with olfactory dysfunction at baseline had lower olfactometric scores than normally smelling individuals (mean [CI95], 8.67 [7.73, 9.60] vs. 10.58 [10.36,10.81], 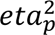=0.19, *p*<0.005) and the same applied for participants impaired at follow-up (mean [CI95], 8.40 [7.33, 9.47] vs. 10.56 [10.34, 10.78], 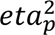=0.22, *p*<0.005).

### Olfactory bulb volume and olfactory function

The mean OB volume was 45.53 [CI95, 43.84, 47.21] mm³. The ANCOVA yielded no significant group difference in OB volume at acute infection between individuals with and without olfactory dysfunction (mean [CI95], 45.85 [43.26, 48.43] vs. 45.21[40.83, 49.59], 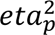=0.00, *p*=0.837; *figure 2b*). Individuals with self-reported sustained olfactory dysfunction at baseline and at follow-up had significantly lower OB volume than normally smelling subjects at that time (mean [CI95], baseline: 40.76 [36.08, 45.44] vs. 46.74 [44.22, 49.26], 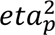=0.03, *p*=0.046; follow-up: 40.45 [35.51, 45.38] vs. 46.55 [44.07, 49.03], 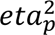=0.03, *p*=0.036). In participants with olfactory dysfunction lower OB volume was accompanied by a shorter time-period between a positive PCR for SARS-CoV-2, signifying the timepoint of acute SARS-CoV-2 infection, and the baseline examination (*r*_*sp*_=0.161, *p*=0.016; *figure 2c*).

### Longitudinal prediction of olfactory function

OB volume derived from MRI at baseline was significantly linearly associated with olfactometry scoring at follow-up (*r*_*sp*_=0.154, *p*=0.025; *figure 2d*). Participants with sustained olfactory dysfunction at follow-up had lower OB volume at baseline (mean [CI95], 40.64 [35.51, 45.77] vs. 47.58 [44.68, 50.48], 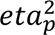=0.06, *p*=0.018; *figure 2e*) than those that had recovered by the time of reassessment.

### Clinical analysis

To further explore potential clinical implications of our findings, correlations of OB volume and neuropsychological cognitive test scores and psychiatric test scores were performed. To summarize, no significant associations of OB volume and scores of the Trail Making Test B (*r*_*sp*_=0.014, *p*=0.722), Animal Naming Test (*r*_*sp*_=-0.090, *p*=0.069), Word List Recall Test (*r*_*sp*_=-0.050, *p*=0.587), Mini Mental State Exam (*r*_*sp*_=-0.025, *p*=0.689), PHQ-9 (*r*_*sp*_=0.098, *p*=0.404) and EQ-5D (*r*_*sp*_=-0.009, *p*=0.306) were found. Corresponding visualizations are displayed in *supplementary materials E7*.

## DISCUSSION

We report on an association of OB volume and olfactory dysfunction in a large sample of mainly mildly to moderately affected COVID-19 convalescents. Longitudinal assessment demonstrated sustained olfactory dysfunction up to two years after acute infection. Participants suffering from olfactory dysfunction beyond acute infection had a significantly lower OB volume at baseline than normally smelling individuals. Moreover, OB volume was predictive for olfactometric performance in the Sniffin’ Sticks test 22 months after the acute infection as well as for the binary outcome of recovery from olfactory dysfunction at follow-up. Neuropsychological test performances were not significantly associated with OB volume. Taken together, our findings suggest that lower volume of the OB may be a promising surrogate marker of smelling function in COVID-19 at post-acute disease stages.

### Longitudinal trajectory of olfactory dysfunction

Its frequency and the concomitant effects on quality of life render olfactory dysfunction a burdensome symptom of COVID-19. Our 2-year longitudinal assessment of olfactory function provided insights about time-dependent development of these symptoms. At acute infection, the proportion of participants reporting olfactory dysfunction was 67.1% which is coherent with previous study reports ranging from 30% to 85%.^(1–4, 6, 7)^ After the acute infection, the prevalence decreased to 21% at the baseline examination (on average 253 days post infection) and 17.5% at follow-up (on average 664 days post infection; *figure 2a*). These numbers support previous literature finding high rates of long term olfactory dysfunction in COVID-19.^(43–45)^ Of note, a recent meta-analysis reports persistent olfactory dysfunction in less patients (11.6%; 95% confidence interval 5.2% to 23.9%).^(46)^ We attribute this difference to design differences with respect to our work: the studies included in the meta-analysis also considered young adults (>18 years) possibly exhibiting higher regenerative capacity as well as recent SARS-CoV-2 variants demonstrably affecting patient olfaction less severely than the wild type variant mainly prevalent at the pandemic onset.^(47)^ Overall, these findings indicate that although most former COVID-19 patients completely recover, subjective olfactory dysfunction persisted in relevant proportion of individuals.

To assess long term olfactory outcomes, olfactometry with the Sniffin’ Sticks Screening 12 test was performed at follow-up. Participants suffering from olfactory dysfunction beyond the acute infection, plausibly showed significantly lower olfactometry scores than normally smelling or recovering subjects. Collectively, our results support previous evidence of sustained olfactory dysfunction in around 20% of patients underscoring the possible long lasting burden following a SARS-CoV-2 infection.^(6, 44)^

### Long-term olfactory dysfunction is associated with lower olfactory bulb volume

Olfactory dysfunction is demonstrably accompanied by lower OB volume in many otorhinolaryngological conditions like post-infectious olfactory disorder, head trauma as well as acute and chronic rhinosinusitis.^(18)^ In line with this notion, our work showed a lower OB volume in individuals with sustained olfactory dysfunction after the acute SARS-CoV-2 infection indicating COVID-19-related OB atrophy (*figure 2b*). Thus, our results corroborate previous reports derived from case studies and small samples suggesting lower OB volume in SARS-CoV-2-induced olfactory dysfunction.^(23–26)^

Previous reports demonstrated an inverse correlation between OB volume and the duration of symptoms in post-infectious olfactory disorder suggesting its predictive capacity.^(22)^ To further investigate this, we related the OB volume with olfactometry scores which were acquired approximately 1 year after the MRI. Notably, the OB volume predicted olfactometry scores at follow-up (*figure 2d*). Yet, the observed correspondence was of a rather low degree, indicating that further determinants of long-term olfactory dysfunction should be considered. Possibly, MRI assessment closer to the acute infection would have resulted in the observation of stronger effects. Furthermore, OB volume was higher in participants in which the smelling sense fully recovered until follow-up compared to those with sustained olfactory dysfunction (*figure 2e*). Hence, the OB volume appears to be a predictor of recovery from olfactory dysfunction, i.e., potentially capturing the severity of damage SARS-CoV-2 exerts on the olfactory system. Interestingly, individuals exhibited higher OB volume the longer the time interval between the positive PCR and the MRI was (*figure 2c*). This might indicate that an increasing OB volume may reflect recovery of olfactory function. However, longitudinal imaging assessment is warranted here. Taken together, these findings suggest that a lower OB volume indicates more severe disruption of the olfactory system and predict persistent olfactory dysfunction in COVID-19 patients.

### Pathomechanistic underpinnings of abnormal olfactory bulb volume

There are multiple potential mechanisms that might explain the observed link between olfactory dysfunction and OB dysintegrity. In the olfactory mucosa, olfactory sensory neurons (OSN) – sensing molecular signatures as odor information – as well as supporting epithelial cells (sustentacular cells) ensure proper sense of smell. Sustentacular cells express angiotensin-converting enzyme 2 (ACE-2) and appear to be a major infection target of the virus.^(13, 48)^ As they support OSN in a glial-like fashion, impairment of sustentacular cells is considered to contribute to COVID-19-related olfactory dysfunction.^(49)^ How OSN are affected by SARS-CoV-2 is controversial. Discussed mechanisms are neurotropism, affection by an impaired support system and damage caused by the immune response to the virus.^(13, 49, 50)^ Recent analyses failed to detect signs of neurotropism and neural invasion through SARS-CoV-2 challenging the notion of the olfactory system serving the virus as an entry point.^(49, 51, 52)^ As the OB serves as a relay for projections from the OSN, volumetric reductions might occur as an effect of indirect OSN affection - e.g., via inflammation or microvasculopathy – rather than direct damage from the virus leading to reduced tissue integrity.^(52)^ Taking the link between OB integrity and long-term olfactory function into account, OB volume might serve as an indicator of severe structural disruption of the olfactory system which corresponds with unfavorable outcomes. Nonetheless, further longitudinal neuroimaging research is warranted to support this notion.

### Olfactory bulb volume is not associated with neuropsychological test scores

By now, COVID-19 is recognized to cause post-acute neurological and psychiatric symptoms like executive dysfunction, fatigue, anxiety, depression and sleep impairment.^(53–56)^ Coherent with these observations, a comprehensive MRI analysis on COVID-19 convalescents from the UK Biobank has shown widespread gray matter volume reductions in areas receiving projections from the olfactory cortex.^(54)^ Its evident exposition to deleterious SARS-CoV-2 effects renders the OB a candidate indicator of COVID-19-related neuropathology beyond the olfactory system. Thus, we tested whether the OB volume is associated with results of neuropsychological test scores. OB volume showed no significant association with tests of cognitive function (Trail Making Test B, Word List Recall, Animal Naming Test), depressive symptoms (Patient Health Questionnaire-9, PHQ-9) and quality of life (EQ-5D). Consequently, pathology of the olfactory system might be disjunct to non-olfaction-related neuropathology in SARS-CoV-2 infection. Nonetheless, our findings might be partially attributable to the overall mild to moderate disease course captured in our sample resulting in negative results. Further investigations of determinants of neurological and psychiatric sequelae of COVID-19 are necessary.

### Strengths and limitations

The strengths of this work lie in its considerable sample size; high quality imaging and phenotypical data; a modern fully-automated MRI-based segmentation of the OB enabling volumetry at scale; olfactory assessment at different time points post infection, including quantitative olfactory testing (at follow-up) providing longitudinal information for up to 2 years. Yet, this study has some limitations. First, MRI acquisition and olfactometry with Sniffin’ sticks were performed only at one timepoint, which makes it difficult to completely address pre-infectious group differences. For instance, our results could partially be explained by individuals with lower OB volume being more susceptible to olfactory dysfunction caused by SARS-CoV-2 infection. Previous work hypothesizes that smaller OB volume and pre-existing reduced number of olfactory receptor neurons increased the patient’s vulnerability to develop post-infectious olfactory loss. With less functional tissue existing in the first place, damage to existing sensory cells might lead to more pronounced olfactory dysfunction.^(57)^ Here, evidence from future longitudinal studies is warranted. As more severely impaired individuals might be more motivated and thus more likely to participate in our study than the average population, our results could possibly be influenced by our recruitment strategy. Additionally, a more in-depth assessment of olfactory function based on odor thresholds was not conducted at follow-up because high dropout rates due to compliance issues were expected. Parosmia may serve as a potential confounding factor in the observed associations, thus necessitating cautious interpretation of our results. Unfortunately, information on parosmia was not available for this study. Lastly, the different SARS-CoV-2 strains appear to differ in terms of olfactory dysfunction frequency and intensity. Our study lacks information about SARS-CoV-2 strains rendering it incapable to address inter-strain differences. However, the investigation started at an early stage of the pandemic most likely soothing the problem of different COVID-19 strains and vaccinations as confounders.

## Conclusion

In this work, we performed OB volumetry as a neuroimaging marker of olfactory dysfunction in patients recovered from mainly mild to moderate COVID-19. By revealing an alteration of the OB in participants with olfactory dysfunction, our results highlight the relevance of the olfactory system in the overall pathophysiology of the disease. However, a connection between OB volume and neuropsychological signs of COVID-19 could not be established. Collectively, these results demonstrate that the OB is a promising target for assessment of olfactory dysfunction in COVID-19.

## ACKNOWLEDGEMENTS

The authors wish to acknowledge all participants of the Hamburg City Health Study and cooperation partners, patrons and the Deanery from the University Medical Center Hamburg—Eppendorf for supporting the Hamburg City Health Study. Special thanks applies to the staff at the Epidemiological Study Center for conducting the study. The participating institutes and departments from the University Medical Center Hamburg-Eppendorf contribute all with individual and scaled budgets to the overall funding. The Hamburg City Health Study is also supported by Amgen, Astra Zeneca, Bayer, BASF, Deutsche Gesetzliche Unfallversicherung (DGUV), DIFE, the Innovative medicine initiative (IMI) under grant number No. 116074 and the Fondation Leducq under grant number 16 CVD 03., Novartis, Pfizer, Schiller, Siemens, Unilever and “Förderverein zur Förderung der HCHS e.V.”. The publication has been approved by the Steering Board of the Hamburg City Health Study.

## AUTHORSHIP CONTRIBUTION

We describe contributions to the paper using the CRediT contributor role taxonomy. Conceptualization: M.P, A.S.H.; Data Curation: M.P., B.B., M.S., C.M., F.N., A.S.H.; Formal analysis: M.P.; Funding acquisition: G.T, B.C, C.B., A.S.H.; Investigation: M.P., B.B., M.S., C.M., F.N., E.P., R.T., G.T., B.C., C.B., A.S.H.; Methodology: M.P.; Software: M.P.; Supervision: C.B., A.S.H.; Visualization: M.P.; Writing—original draft: M.P., B.B.; Writing—review & editing: M.P., B.B., M.S., C.M., F.N., E.P., R.T., G.T., B.C., C.B., A.S.H. Data access: M.P., B.B., and A.S.H. had full access to all the data in the study and take responsibility for the integrity of the data and the accuracy of the data analysis.

## DATA AVAILABILITY

Data will be made available on reasonable request from any qualified investigator after evaluation of the request by the Steering Board of the HCHS. Analysis code and documentation is publicly available on GitHub (https://github.com/csi-hamburg/CSIframe/blob/main/pipelines/obseg/obseg.sh, https://github.com/csi-hamburg/CSIframe/wiki/Olfactory-bulb-segmentation and https://github.com/csi-hamburg/2022_petersen_ob_postcovid).

