## Supplementals for "Reduced Olfactory Bulb Volume Accompanies Olfactory Dysfunction After Mild SARS-CoV-2 Infection"

#### E1 – Investigation timeline

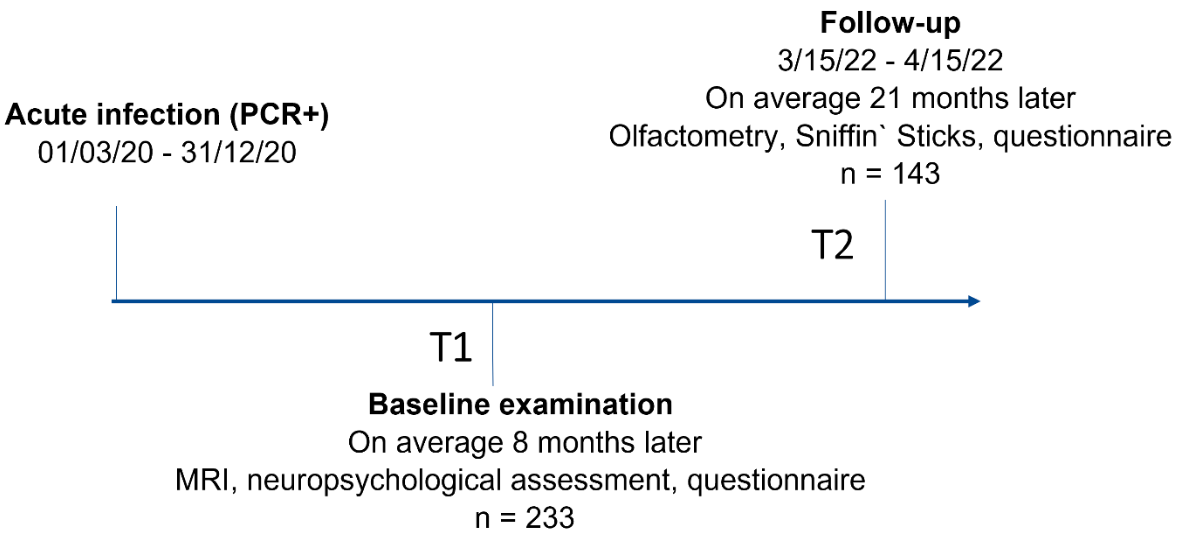

#### E2 - Exemplary visualizations of the olfactory bulb – First subject (1)

First subject – no subjective olfactory dysfunction, male, age range 56-60 y., olfactory bulb volume: 67.68 mm³

Overview of the segmentations of the olfactory bulb. Panel from left to right: coronal, axial, sagittal slices (blue: left olfactory bulb, red: right olfactory bulb)

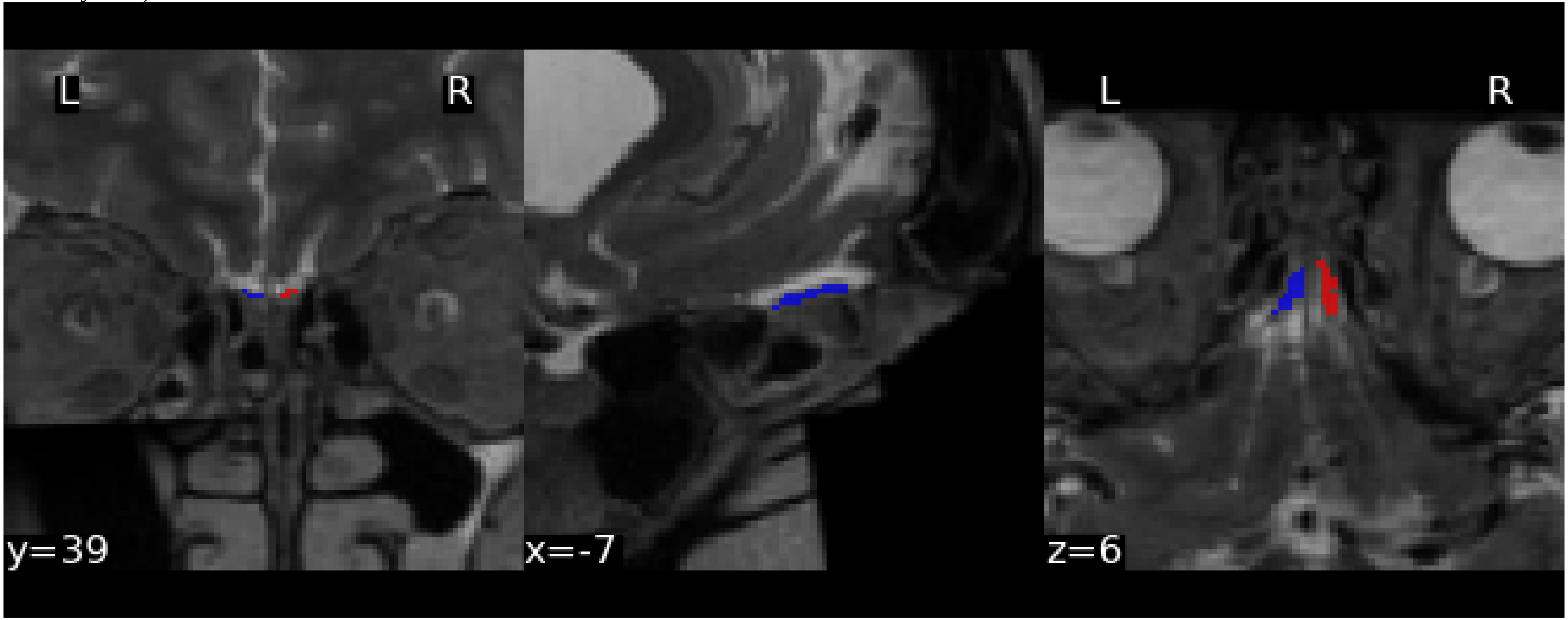

### E3 - Exemplary visualizations of the olfactory bulb – First subject (2)

Coronal slices in lightbox view. Left panel without segmentations, right panel with segmentations

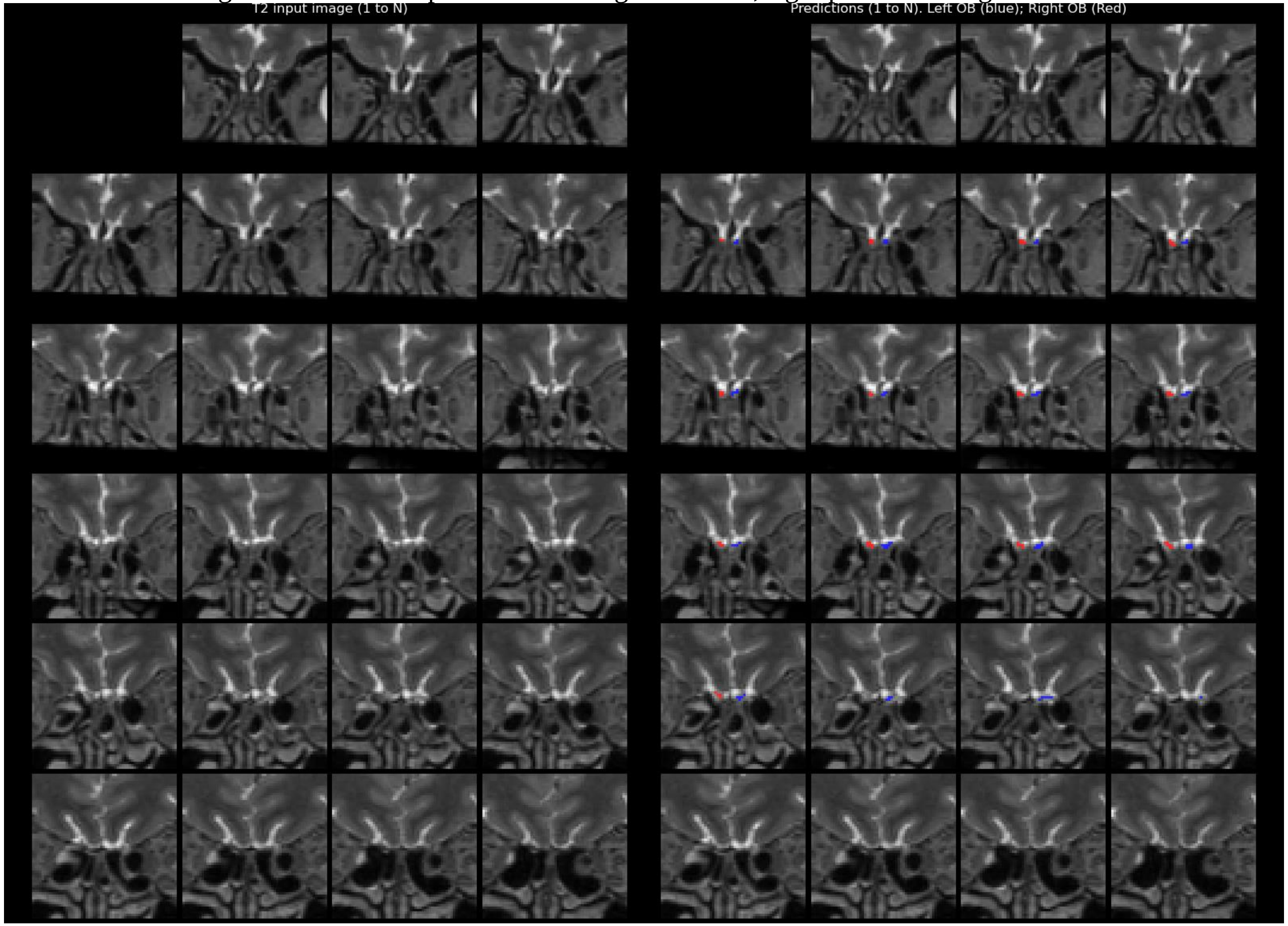

#### E4 - Exemplary visualizations of the olfactory bulb – Second subject (1)

Second subject – with subjective olfactory dysfunction, male, age range 56-60 years, olfactory bulb volume: 28.67 $mm^3$

Overview of the segmentations of the olfactory bulb. Panel from left to right: coronal, axial, sagittal slices (blue: left olfactory bulb, red: right olfactory bulb)

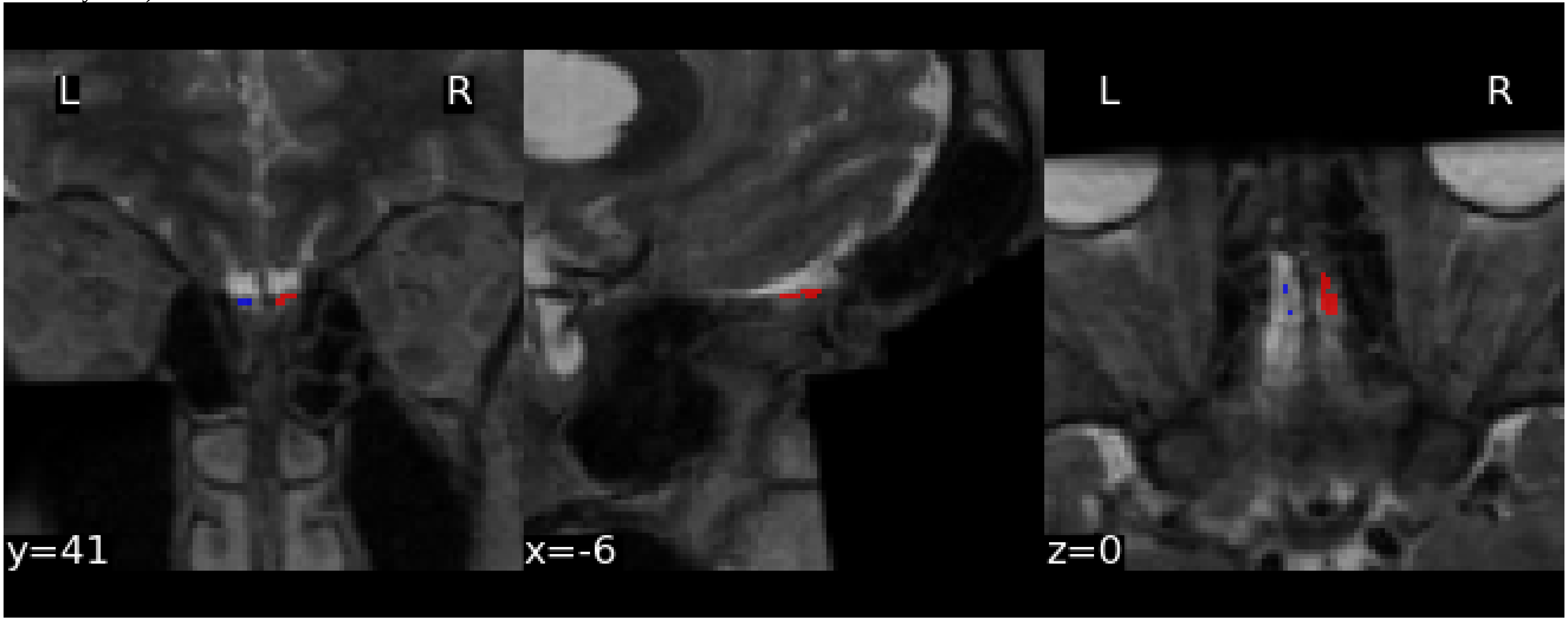

### E5 - Exemplary visualizations of the olfactory bulb – Second subject (2)

Coronal slices in lightbox view. Left panel without segmentations, right panel with segmentations

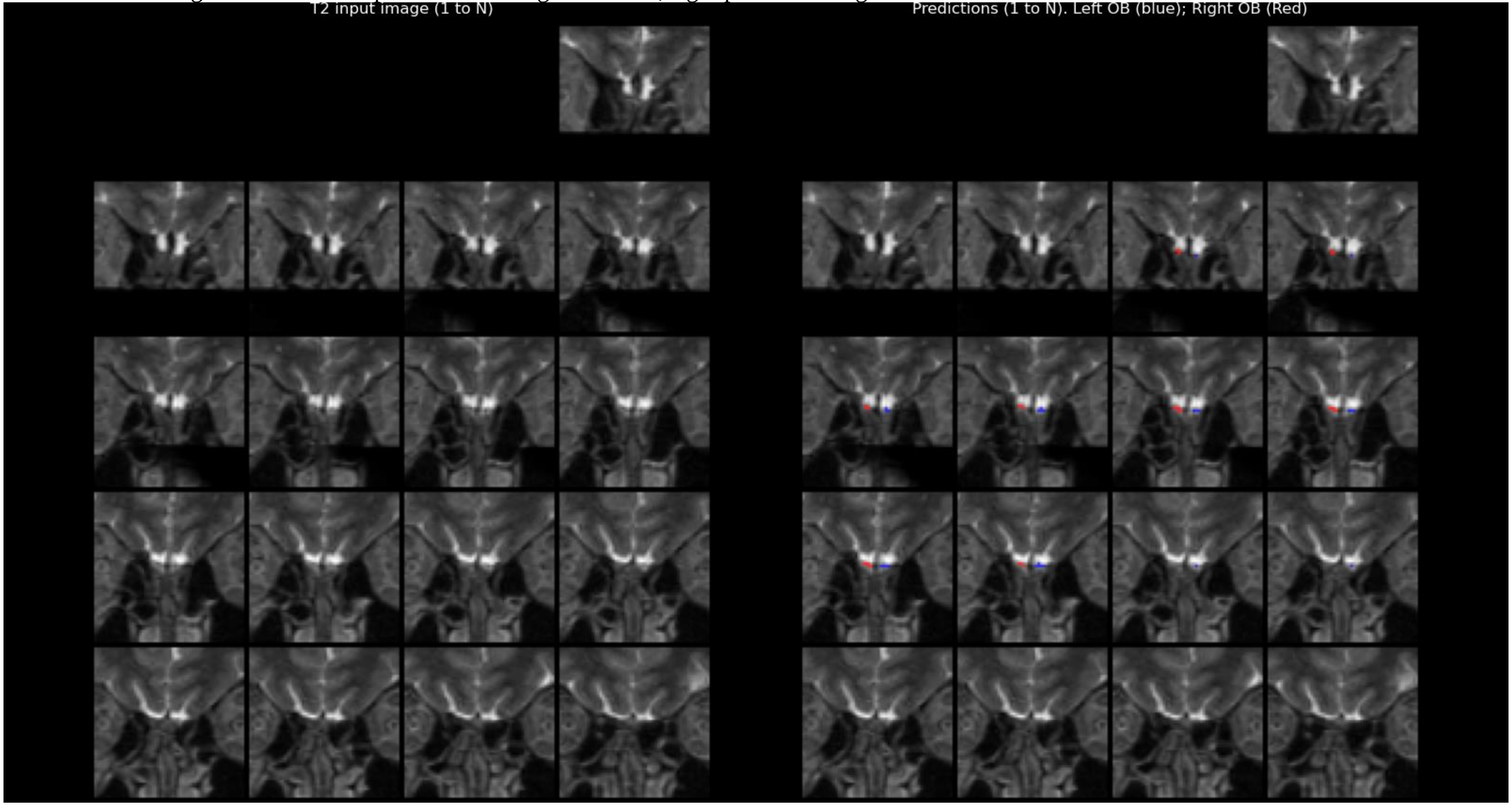

### E6 – Group comparison olfactometry scores

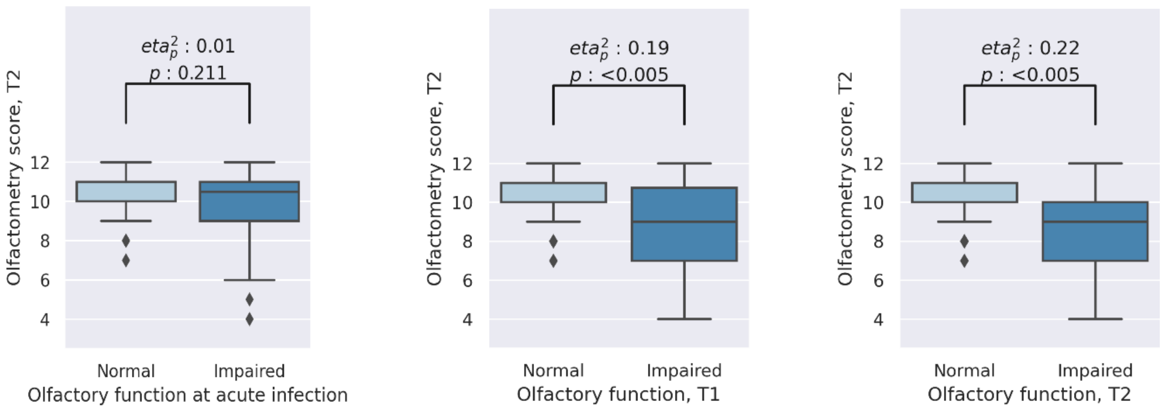

### E7 – Linear associations olfactory bulb volume and neuropsychological test performances

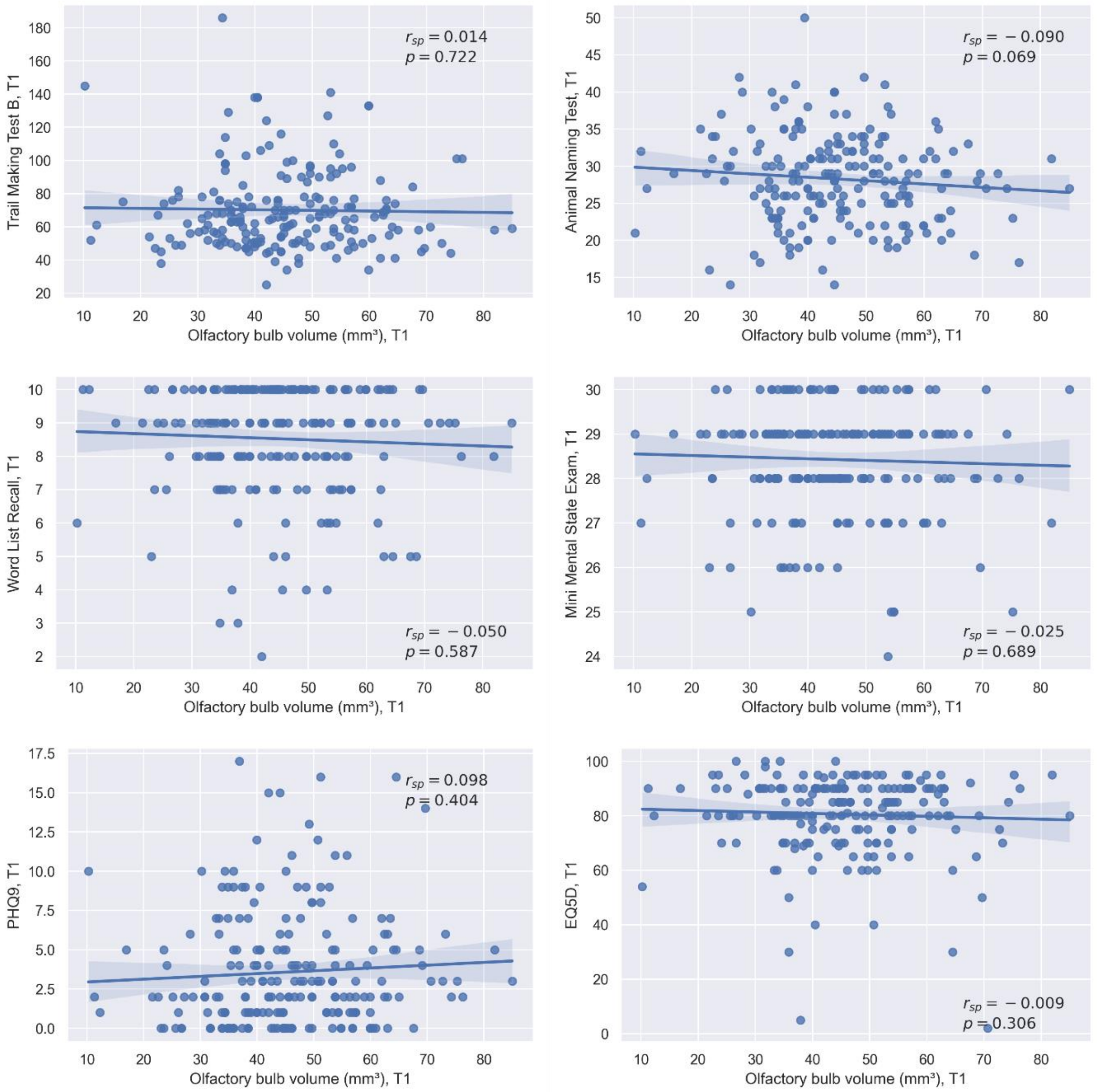


Linear associations between olfactory bulb volume and the Trail Making Test B, Word List Recall, Animal Naming Test, Mini Mental State Exam, Patient Health Questionnaire 9 and EQ-5D are shown. No significant relationships were found. Abbreviations: p = p-value, PHQ-9 = Patient Health Questionnaire 9, $r_{sp}$ = spearman correlation coefficient, T1 = at baseline, T2 = at follow-up.
